# A novel immunotherapy prognostic score for patients with pretreated advanced urinary tract carcinoma from the subgroup analysis of the SAUL study. The urInary TrAct CArcinoma score (ITACA)

**DOI:** 10.1101/2022.08.30.22279352

**Authors:** Giuseppe Fornarini, Sara Elena Rebuzzi, Sebastiano Buti, Pasquale Rescigno, Axel Merseburger, Ugo de Giorgi, Umberto Basso, Marco Maruzzo, Patrizia Giannatempo, Marta Ponzano, Emilio Francesco Giunta, Fabio Catalano, Veronica Murianni, Alessandra Damassi, Malvina Cremante, Annalice Gandini, Silvia Puglisi, Miguel Angel Llaja Obispo, Alessio Signori, Giuseppe Luigi Banna

## Abstract

**Background:** The current prognostic models for patients with advanced urinary tract cancers were developed and validated in the chemotherapy setting. As immunotherapy has become the backbone of novel treatments, updated prognostic scores are needed.

**Methods:** A comprehensive analysis of inflammatory indexes from peripheral blood and clinical factors was planned on the entire real-world cohort of pretreated patients with advanced urinary tract carcinoma receiving atezolizumab in the prospective, single-arm, phase IIIb SAUL study. Univariable and multivariable analyses with overall survival as the primary endpoint, bootstrap internal validation, Schneeweiss scoring system and calibration test were performed to develop a novel immunotherapy prognostic score.

**Results:** Thirteen clinical variables from 1001 patients were analysed. The following eight prognostic factors were included in a model: ECOG PS, liver and bone metastases, histology, pre-treatment steroids, systemic immune-inflammatory index (i.e., neutrophils-to-lymphocytes ratio times platelets count), haemoglobin and lactate dehydrogenase.

The prognostic model was able to stratify patients into five risk groups with significantly different (p < 0.001) median overall survival of NR, 18.0, 8.7, 4.6 and 2.4 months, respectively. The c-index for OS was higher than the Bellmunt score one (0.702 vs 0.672).

**Conclusions:** A novel 5-class prognostic model contemporary to immunotherapy provides robust prognostic discrimination of patients with advanced urinary tract carcinoma homogeneously treated with immunotherapy through baseline affordable and reproducible clinical and laboratory factors. It couls be quickly adopted in clinical practice to inform patients about prognosis with immunotherapy and assess the benefit of novel immunotherapy combinations in clinical trials.

## Introduction

In recent years, the European Medicines Agency (EMA) has approved immune checkpoint inhibitors (ICI) targeting the programmed cell death protein-1 (PD-1)/ ligand-1 (PD-L1) as second-line treatments for advanced urothelial carcinoma (aUC) after platinum-based chemotherapy (pembrolizumab, nivolumab and avelumab), as first-line in platinum-ineligible patients with high PD-L1 expressing tumours (atezolizumab and pembrolizumab), or as maintenance following platinum-based chemotherapy (avelumab)^1-5^.

Despite this, patients’ stratification and treatment choices do not rely on robust predictive biomarkers. At the same time, available prognostic models were developed and validated when treatment options for aUCs did not include immunotherapies^6^.

SAUL was a large real-world phase IIIb prospective study evaluating atezolizumab in patients with advanced urothelial or nonurothelial urinary tract carcinoma progressing during or after one to three prior therapies^7^. In a sub-analysis of this study on the Italian cohort, we postulated that the combination of the systemic immune □ inflammation index (SII = neutrophil-to-lymphocyte ratio [NLR] x platelet count) and PDL-1 expression could stratify patients into three risk categories correlated with progression-free (PFS) and overall survivals (OS)^8^.

In the current work, we present a more comprehensive analysis of inflammatory indexes, tumour’s and patients’ characteristics in the SAUL overall population leading to a novel prognostic score to predict survival and identify patients likely to benefit from immunotherapy.

## Materials and Methods

The sponsor provided the complete SAUL database after approval of the subanalysis project (with data cut-off on 16^th^ September 2018) through the VIVLI Global Clinical Trials Data Sharing Platform. The trial was approved by each participating centre’s institutional review board or ethics committee, and all patients provided written informed consent.

### Study design and study population

The SAUL (NCT02928406) was a single-arm open-label phase IIIb safety study of atezolizumab in a real-world population of patients with locally advanced or metastatic urothelial or nonurothelial carcinoma of the urinary tract. A detailed description of the study design and the main results on 1004 enrolled patients have already been published^7^.

All participants were required to have Eastern Cooperative Oncology Group (ECOG) Performance Status (PS) ≤ 2 and disease progression during or following 1-3 prior platinum- or non-platinum-based treatments. Patients with renal impairment, controlled central nervous system (CNS) metastases or autoimmune disease, and concomitant corticosteroids were eligible.

Atezolizumab 1200 mg was administered intravenously every three weeks until lack of clinical benefit, unacceptable toxicity, patient’s or investigator’s decision to discontinue therapy, or death. Radiological assessments were carried out every nine weeks for 12 months and then every 12 weeks. Immunohistochemical assessment of PD-L1 expression was performed on tumour specimens using the VENTANA SP142 antibody.

### Prognostic factors

Baseline clinical variables included patients’ characteristics (ECOG PS, body mass index [BMI], pretreatment steroids, number of previous therapy lines), tumour features (histology, PD-L1 expression, liver and bone metastases) and laboratory data (haemoglobin and albumin levels, NLR, SII, lactate dehydrogenase [LDH]).

### Study endpoints

The primary endpoint for this analysis was developing a prognostic score for immunotherapy based on independent prognostic factors on OS. The OS was calculated from first atezolizumab administration until death, censored at last follow-up for patients who were alive. The prediction of the PFS was a secondary endpoint. The PFS was calculated from first atezolizumab administration until first radiographic/clinical progression or death, whichever occurred first, censored at last follow-up for patients who were alive without progression.

### Statistical methods

Patients’ characteristics were presented using absolute frequency and percentage for categorical variables and by mean with standard deviation or median and range for quantitative variables.

Univariable association between each covariate and clinical outcome (OS, or PFS) was assessed by the Cox proportional hazards model. The Kaplan-Meier (KM) method was used to estimate the survival curves of OS and PFS.

Cut-offs for categorical variables were chosen according to survival Receiver Operating Characteristic (ROC) curves or predetermined cut-offs where available. As non-univocal cut-offs are available for NLR and SII, the two most reported cut-offs in the literature (3 and 5 for NLR; 1375 and 884 for SII)^8-9^ and the ROC-identified cut-offs (6.2 for NLR and 1589 for SII) were used.

All variables with a p-value < 0.10 at the univariable analysis were selected for inclusion in the multivariable model. Only those variables that resulted as statistically significant with a p-value < 0.05 at the multivariable analysis were retained for the prognostic model.

If both the SII and NLR were found suitable for inclusion in the multivariable model, their simultaneous inclusion in the model would be avoided as the SII entails the NLR. In that case, we planned to perform two different multivariable models with NLR and SII, and to consider only the model with the highest Harrell’s c-index for developing the prognostic score.

The variable selection process for the prognostic score creation and the parameter estimation from the Cox models were internally validated by generating 500 bootstrap samples with replacement. Variables that remained in more than 50% of the bootstrap samples were selected for inclusion in the final prognostic score.

A bias-corrected estimate of the discriminatory ability (c-index) was calculated using 500 bootstrap samples (R rms package) to avoid overfitting during the building and estimation of the prognostic score.

The regression coefficient-based (Schneeweiss) scoring system was adopted to assign a weight to each factor included in the prognostic score^11^. The weights of each of those factors were obtained from the coefficients of the Cox regression model.

Finally, the prognostic score was stratified in risk strata according to the likelihood-ratio test and after checking the survival estimates of the score.

An exploratory comparison of the bootstrapped score’s c-indexes for OS and PFS was performed with Bellmunt’s ones.

Calibration of the prognostic score was assessed at six months and 12 months. Bootstrap with 300 replications was used to get overfitting-corrected estimates of predicted survival probability that were compared with observed survival estimates from the KM method.

The clinical utility of the prognostic score was explored using the decision-curve analysis (DCA). The net benefit reported in the DCA helps determine whether or in which circumstances using the prognostic score for clinical decisions results in more good than harm.

Since neutrophils, lymphocytes, platelets, LDH showed a variable quote of missing values, multiple imputations (N = 10 imputations) with chained equations algorithm were used to estimate the sets of plausible values. Each imputation model included the variable with missing data as the dependent variable and variables with complete data (ECOG PS, age, gender, tobacco history, previous lines of therapy) as independent variables.

Stata (v.17; StataCorp LLC) and R (v.4.1.1) were used for the computation.

## Results

### Patients’ characteristics

Overall, 1004 patients were enrolled on the SAUL study. Of these, data of 1001 (99.7%) were available in the database provided by the sponsor. All the 1001 patients were eligible for inclusion in this analysis. Their clinical characteristics are summarised in Table 1. The majority of the patients were male (77.7%), presented with an ECOG PS of 0-1 (90.2%), had urothelial histology (95.3%), did not have liver or bone (respectively, 72.6% and 75.9%) metastases and received less than two lines of therapy (61.7%). The median age of the overall cohort was 68 years (range, 60-74).

**Table 1.** Patients’ characteristics.

| Characteristics | N (%) |
| --- | --- |
| Age years, median (IQR) | 68 (60-74) |
| Sex |  |
| Male | 778 (77.7) |
| Female | 223 (22.3) |
| BMI |  |
| Median (IQR) | 25.7 (23.1-29.1) [N = 990] |
| <25 | 428 (42.8) |
| ≥25 | 562 (56.1) |
| Missing | 11 (1.1) |
| ECOG PS |  |
| 0 | 427 (42.7) |
| 1 | 476 (47.5) |
| 2 | 98 (9.8) |
| PD-L1 expression |  |
| IC0 | 243 (24.3) |
| IC1 | 420 (42.0) |
| IC2 | 201 (20.0) |
| IC3 | 67 (6.7) |
| Missing | 70 (7.0) |
| PD-L1 expression, groups |  |
| IC0-1 | 663 (66.3) |
| IC2-3 | 268 (26.7) |
| Missing | 70 (7.0) |
| Histological type |  |
| Urothelial | 954 (95.3) |
| Nonurothelial | 35 (3.5) |
| Mixed | 12 (1.2) |
| Histology grade |  |
| 1 | 54 (5.4) |
| 2 | 118 (11.8) |
| 3 | 782 (78.1) |
| Missing | 47 (4.7) |
| Tumor location |  |
| Bladder | 747 (74.6) |
| Renal pelvis | 122 (12.2) |
| Ureter | 98 (9.8) |
| Urethra | 10 (1.0) |
| Other | 24 (2.4) |
| Regional lymph node at study entry |  |
| N0 | 308 (30.8) |
| N1 | 215 (21.4) |
| N2 | 247 (24.7) |
| N3 | 172 (17.2) |
| Not applicable | 59 (5.9) |
| Distant metastasis at study entry |  |
| M0 | 61 (6.1) |
| M1 | 940 (93.9) |
| Liver metastasis at study entry |  |
| No | 727 (72.6) |
| Yes | 274 (27.4) |
| Bone metastasis at study entry |  |
| No | 760 (75.9) |
| Yes | 241 (24.1) |
| Previous lines of cancer therapy |  |
| 0 | 383 (38.3) |
| 1 | 546 (54.5) |
| 2-3 | 72 (7.2) |
| LDH at study entry, median (IQR) | 238 (184-352) [N = 963] |
| Neutrophils at study entry, median (IQR) | 5.1 (3.7-7.0) [N = 777] |
| Lymphocytes at study entry, median (IQR) | 1.3 (1-1.8) [N = 769] |
| Platelets, median (IQR) | 248 (196-317) [N = 993] |
| NLR, median (IQR) | 3.79 (2.50-5.90) [n=769] |
| SII, median (IQR) | 923.8 (586.5-1591.1) [n=769] |
| LDH at study entry imputed, median (IQR) | 242 (185-354) |
| Neutrophils at study entry imputed, median (IQR) | 5.0 (3.8-6.7) |
| Lymphocytes at study entry imputed, median (IQR) | 1.4 (1.1-1.7) |
| Platelets imputed, median (IQR) | 248 (196-317) |
| NLR imputed, median (IQR) | 3.82 (2.67-5.55) |
| SII imputed, median (IQR) | 923.8 (597.7-1582.8) |

Abbreviations: BMI, body mass index; ECOG PS, Eastern Cooperative Oncology Group performance status; IC immune cells; IQR, interquartile range; LDH, lactate dehydrogenase; N, number of patients; NLR, neutrophils-to-lymphocytes ratio; PD-L1, programmed death-ligand 1; SII, systemic immune-inflammatory index (i.e., NLR times platelets count).

Imputed data for missing ones were consistent with those with complete information. At the data cut-off for the primary analysis, after a median follow-up of 6.8 months (interquartile range [IQR]: 3.0-11.1), 728 patients (72.7%) progressed, and 553 patients (55.2%) died.Median OS was 8.7 months (95% CI 7.8-9.9), median PFS was 2.2 months (95% CI 2.1-2.4).

### Univariable analyses for OS

Thirteen clinical variables were studied for their correlation with OS by the univariable analysis. Related median OS (mOS), hazard ratio (HR) with 95% confidence interval (CI), p-value and c-index are presented in Table 2. All variables except the number of prior therapy lines (p = 0.64) were statistically significantly associated with OS (p<0.05), including the well-known prognostic factors for aUC (e.g. ECOG PS, visceral metastases, haemoglobin level). The highest c-index for the NLR and SII was found with the cut-offs of 5 and 884, respectively.

**Table 2.** Univariable and multivariable Cox regression analyses on OS (N = 1001).

| Characteristics | N (%) | mOS | Univariable |  |  | Multivariable NLR<br>(c-index = 0.717) |  | Multivariable SII<br>(c-index = 0.723) |  |
| --- | --- | --- | --- | --- | --- | --- | --- | --- | --- |
|  |  |  | HR (95% CI) | p value | c-index | HR (95% CI) | p value | HR (95% CI) | p value |
| **NLR** (continuous) |  |  | 1.10 (1.08-1.11) | <0.001 | 0.619 |  |  |  |  |
| <3 | 331 (33.1) | 12.7 | 1.00 (ref) |  | 0.562 |  |  |  |  |
| ≥3 | 670 (66.9) | 7.1 | 1.66 (1.37-2.00) | <0.001 |  |  |  |  |  |
| <5 | 688 (68.7) | 10.9 | 1.00 (ref) |  | 0.594 | 1.00 (ref) |  |  |  |
| ≥5 | 313 (31.3) | 4.4 | 2.17 (1.83-2.57) | <0.001 |  | 1.58 (1.32-1.89) | <0.001 |  |  |
| <6.2 (cut-off found) | 804 (80.3) | 10.6 | 1.00 (ref) |  | 0.583 |  |  |  |  |
| ≥6.2 | 197 (19.7) | 3.6 | 2.58 (2.14-3.12) | <0.001 |  |  |  |  |  |
| **SII** (continuous)<br>(100-unit increase) |  |  | 1.03 (1.02-1.04) | <0.001 | 0.635 |  |  |  |  |
| <1375 | 695 (69.4) | 11.2 | 1.00 (ref) |  |  |  |  |  |  |
| ≥1375 | 306 (30.6) | 4.1 | 2.19 (1.85-2.61) | <0.001 | 0.594 |  |  |  |  |
| <884 | 470 (46.9) | 13.9 | 1.00 (ref) |  |  |  |  | 1.00 (ref) |  |
| ≥884 | 531 (53.1) | 5.5 | 2.16 (1.82-2.57) | <0.001 | 0.602 |  |  | 1.69 (1.41-2.02) | <0.001 |
| <1589 (cut-off found) | 752 (75.1) | 11.2 | 1.00 (ref) |  |  |  |  |  |  |
| ≥1589 | 249 (24.9) | 3.8 | 2.49 (2.09-2.97) | <0.001 | 0.596 |  |  |  |  |
| **PD-L1** |  |  |  |  | 0.534 |  |  |  |  |
| IC0 | 260 (26.0) | 7.2 | 1.00 (ref) |  |  |  |  |  |  |
| IC1 | 470 (47.0) | 8.5 | 0.90 (0.74-1.10) | 0.29 |  |  |  |  |  |
| IC2 | 204 (20.4) | 11.4 | 0.74 (0.58-0.95) | 0.020 |  |  |  |  |  |
| IC3 | 67 (6.7) | NR | 0.64 (0.43-0.95) | 0.026 |  |  |  |  |  |
| **ECOG PS** |  |  |  |  | 0.611 |  |  |  |  |
| 0 | 427 (42.7) | 14.5 | 1.00 (ref) |  |  | 1.00 (ref) |  | 1.00 (ref) |  |
| 1-2 | 574 (57.3) | 5.7 | 2.19 (1.83-2.61) | <0.001 |  | 1.66 (1.38-2.00) | <0.001 | 1.65 (1.37-1.99) | <0.001 |
| **Liver metastases** |  |  |  |  | 0.594 |  |  |  |  |
| No | 727 (72.6) | 11.0 | 1.00 (ref) |  |  | 1.00 (ref) |  | 1.00 (ref) |  |
| Yes | 274 (27.4) | 4.2 | 2.21 (1.86-2.64) | <0.001 |  | 2.14 (1.79-2.55) | <0.001 | 2.07 (1.73-2.47) | <0.001 |
| **Bone metastases** |  |  |  |  | 0.550 |  |  |  |  |
| No | 760 (75.9) | 10.3 | 1.00 (ref) |  |  | 1.00 (ref) |  | 1.00 (ref) |  |
| Yes | 241 (24.1) | 5.8 | 1.70 (1.41-2.04) | <0.001 |  | 1.39 (1.16-1.68) | 0.001 | 1.44 (1.19-1.74) | <0.001 |
| **LDH** (continuous, 100-unit) |  |  | 1.13 (1.10-1.16) | <0.001 | 0.598 |  |  |  |  |
| ≤ULN | 651 (65.0) | 11.2 | 1.00 (ref) |  |  | 1.00 (ref) |  | 1.00 (ref) |  |
| >ULN | 350 (35.0) | 4.8 | 1.95 (1.65-2.31) | <0.001 | 0.587 | 1.64 (1.38-1.95) | <0.001 | 1.61 (1.35-1.91) | <0.001 |
| **Histology** |  |  |  |  | 0.506 |  |  |  |  |
| Urothelial | 954 | 8.9 | 0.69 (0.49-0.98) | 0.036 |  | 1.00 (ref) |  | 1.00 (ref) |  |
| Non-urothelial | 47 | 7.3 | 1.45 (1.03-2.08) | 0.036 |  | 1.47 (1.04-2.08) | 0.031 | 1.52 (1.07-2.16) | 0.018 |
| **BMI** |  |  |  |  | 0.537 |  |  |  |  |
| <25 | 428 | 7.0 | 1.00 (ref) |  |  |  |  |  |  |
| ≥25 | 562 | 10.3 | 0.76 (0.64-0.90) | 0.002 |  |  |  |  |  |
| **Pre-treatment steroids** |  |  |  |  | 0.547 |  |  |  |  |
| No | 782 | 10.0 | 1.00 (ref) |  |  | 1.00 (ref) |  | 1.00 (ref) |  |
| Yes | 219 | 5.0 | 1.67 (1.39-2.01) | <0.001 |  | 1.32 (1.09-1.59) | 0.005 | 1.34 (1.11-1.63) | 0.003 |
| **Previous lines of cancer therapy** |  |  |  |  | 0.504 |  |  |  |  |
| <2 | 929 | 8.8 | 1.00 (ref) |  |  |  |  |  |  |
| ≥2 | 72 | 7.4 | 1.08 (0.77-1.52) | 0.64 |  |  |  |  |  |
| **Hemoglobin level (continuous)** |  |  | 0.97 (0.96-0.98) | <0.001 | 0.655 |  |  |  |  |
| >10 g/dL | 840 (83.9) | 10.0 | 1.00 (ref) |  |  | 1.00 (ref) |  | 1.00 (ref) |  |
| <10 g/dL | 161 (16.1) | 3.8 | 2.15 (1.75-2.65) | <0.001 | 0.559 | 1.47 (1.18-1.83) | 0.001 | 1.50 (1.21-1.87) | <0.001 |
| **Albumin level (continuous)** |  |  | 0.92 (0.90-0.93) | <0.001 | 0.636 |  |  |  |  |
| Normal | 892 (89.1) | 9.5 | 1.00 (ref) |  |  | 1.00 (ref) |  | 1.00 (ref) |  |
| Lower than Normal level | 109 (10.9) | 3.5 | 2.09 (1.65-2.65) | <0.001 | 0.545 | 1.27 (0.99-1.63) | 0.063 | 1.33 (1.04-1.70) | 0.025 |

Abbreviations: BMI, body mass index; ECOG PS, Eastern Cooperative Oncology Group performance status; CI, confidence interval; HR, hazard ratio; IC, immune cells; LDH, lactate dehydrogenase; mOS, median overall survival; N, number of patients; NLR, neutrophils-to-lymphocytes ratio; NR, not reached; OS, overall survival; PD-L1, programmed death-ligand 1; ref, reference; SII, systemic immune-inflammatory index (i.e., NLR times platelets count); ULN, upper limit of normal.

### Multivariable analyses for OS

Ten of the 12 variables from the univariable analysis maintained their statistically significant association with OS (p<0.05) in the multivariable analysis (Table 2). The body mass index (BMI) and PD-L1 tumour expression were the two nonsignificant factors. To welldevelop the prognostic score, we drew two multivariable models based on NLR and SII, including ECOG PS, liver and bone metastases, histology, pretreatment steroids, haemoglobin, albumin and LDH.

### Bootstrap validation and prognostic score development

The multivariable model with SII was preferred to the NLR one for the higher c-index (0.723 vs 0.717). After the internal bootstrap validation, the prognostic score included the following variables remaining in more than 50% of the bootstrap samples: ECOG PS, liver and bone metastases, histology, pretreatment steroids, SII, haemoglobin and LDH (Table 3).

**Table 3.** Bootstrap validation and prognostic score development.

| Characteristics | N | Multivariable after bootstrap selection<br>(c-index corrected for optimism=0.717<br>(95% CI: 0.696-0.738)) |  | Bootstrap (500 replications) |  | Risk-scoring<br>(Schneeweiss) |
| --- | --- | --- | --- | --- | --- | --- |
| | | HR (95% CI) | p-value | β-coefficient ± SE | HR (95% CI) | |
| **SII** |  |  |  |  |  |  |
| <884 | 470 (46.9) | 1.00 (ref) |  | 0 |  | 0 |
| ≥884 | 531 (53.1) | 1.71 (1.43-2.04) | <0.001 | 0.540 ± 0.098 | 1.72 (1.42-2.08) | 2 |
| **ECOG PS** |  |  |  |  |  |  |
| 0 | 427 (42.7) | 1.00 (ref) |  | 0 | 1.00 (ref) | 0 |
| 1-2 | 574 (57.3) | 1.67 (1.39-2.01) | <0.001 | 0.522 ± 0.095 | 1.69 (1.40-2.03) | 2 |
| **Liver metastases** |  |  |  |  |  |  |
| No | 727 (72.6) | 1.00 (ref) |  | 0 | 1.00 (ref) | 0 |
| Yes | 274 (27.4) | 2.07 (1.74-2.48) | <0.001 | 0.745 ± 0.103 | 2.11 (1.72-2.58) | 2 |
| **Bone metastases** |  |  |  |  |  |  |
| No | 760 (75.9) | 1.00 (ref) |  | 0 | 1.00 (ref) | 0 |
| Yes | 241 (24.1) | 1.43 (1.19-1.72) | <0.001 | 0.361 ± 0.101 | 1.43 (1.18-1.75) | 1 |
| **LDH** (continuous, 100-unit) |  |  |  |  |  |  |
| ≤ULN | 651 (65.0) | 1.00 (ref) | | 0 | 1.00 (ref) | 0 |
| >ULN | 350 (35.0) | 1.65 (1.39-1.96) | <0.001 | 0.508 ± 0.100 | 1.66 (1.37-2.02) | 2 |
| **Histology** |  |  |  |  |  |  |
| Urothelial | 954 | 1.00 (ref) |  | 0 | 1.00 (ref) | 0 |
| Non-urothelial | 47 | 1.52 (1.07-2.16) | 0.018 | 0.446 ± 0.189 | 1.56 (1.08-2.26) | 1 |
| **Pre-treatment steroids** |  |  |  |  |  |  |
| No | 782 | 1.00 (ref) |  | 0 | 1.00 (ref) | 0 |
| Yes | 219 | 1.36 (1.12-1.64) | 0.002 | 0.318 ± 0.107 | 1.37 (1.11-1.70) | 1 |
| **HGB level** (continuous) |  |  |  |  |  |  |
| >10 g/dL | 840 (83.9) | 1.00 (ref) |  | 0 | 1.00 (ref) | 0 |
| <10 g/dL | 161 (16.1) | 1.58 (1.28-1.96) | <0.001 | 0.466 ± 0.126 | 1.59 (1.24-2.04) | 2 |

Abbreviations: BMI, body mass index; ECOG PS, Eastern Cooperative Oncology Group performance status; CI, confidence interval; HGB, haemoglobin; HR, hazard ratio; LDH, lactate dehydrogenase; N, number of patients; NLR, neutrophils-to-lymphocytes ratio; ref, reference; SII, systemic immune-inflammatory index (i.e., NLR times platelets count); ULN, upper limit of normal.

The results of the bootstrap analysis were similar to the original SII multivariable model except for albumin (Table 2), suggesting successful internal validation.

According to the regression Schneeweiss coefficient, the prognostic factors with the highest weight (i.e., risk scoring of 2) were SII, ECOG PS, liver metastases, haemoglobin and LDH. The prognostic score stratified patients into five risk groups : patients in group 1 presented with 0-1 risk factors, group 2 with 2-3, group 3 with 4-5, group 4 with 6-7 and group 5 with ≥ 8 factors. Median OS was not reached for group 1, and it was respectively 18 (95% CI: 10.6-25) months, 8.7 (95% CI: 7.3-12.8) months, 4.6 (95% CI: 4.0-6.3) and 2.4 (95% CI: 1.7-2.9) months for the other groups. We observed a statistically significant difference in OS and PFS between these five groups (Table 4 and 1S, Figure 1). Calibration of the risk score showed a good concordance between the predicted and the observed OS at 6 and 12 months (Figure 2). The c-index for OS and PFS of the bootstrapped score resulted higher than the Bellmunt score (for OS: 0.702 vs 0.672, +0.03; for PFS: 0.694 vs 0.613, +0.081) (Tables 2S and 3S, Figure 1S).

**Table 4.**
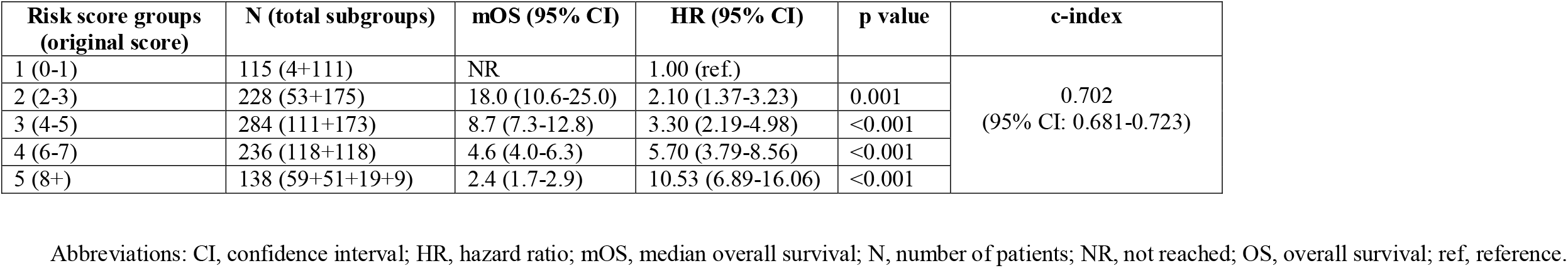
Correlation of the prognostic score with OS.

| **Risk score groups**<br>**(original score)** | **N (total subgroups)** | **mOS (95% CI)** | **HR (95% CI)** | **p value** | **c-index** |
| --- | --- | --- | --- | --- | --- |
| 1 (0-1) | 115 (4+111) | NR | 1.00 (ref.) |  | 0.702<br>(95% CI: 0.681-0.723) |
| 2 (2-3) | 228 (53+175) | 18.0 (10.6-25.0) | 2.10 (1.37-3.23) | 0.001 |  |
| 3 (4-5) | 284 (111+173) | 8.7 (7.3-12.8) | 3.30 (2.19-4.98) | <0.001 |  |
| 4 (6-7) | 236 (118+118) | 4.6 (4.0-6.3) | 5.70 (3.79-8.56) | <0.001 |  |
| 5 (8+) | 138 (59+51+19+9) | 2.4 (1.7-2.9) | 10.53 (6.89-16.06) | <0.001 |  |

Abbreviations: CI, confidence interval; HR, hazard ratio; mOS, median overall survival; N, number of patients; NR, not reached; OS, overall survival; ref, reference.

**Figure 1.**
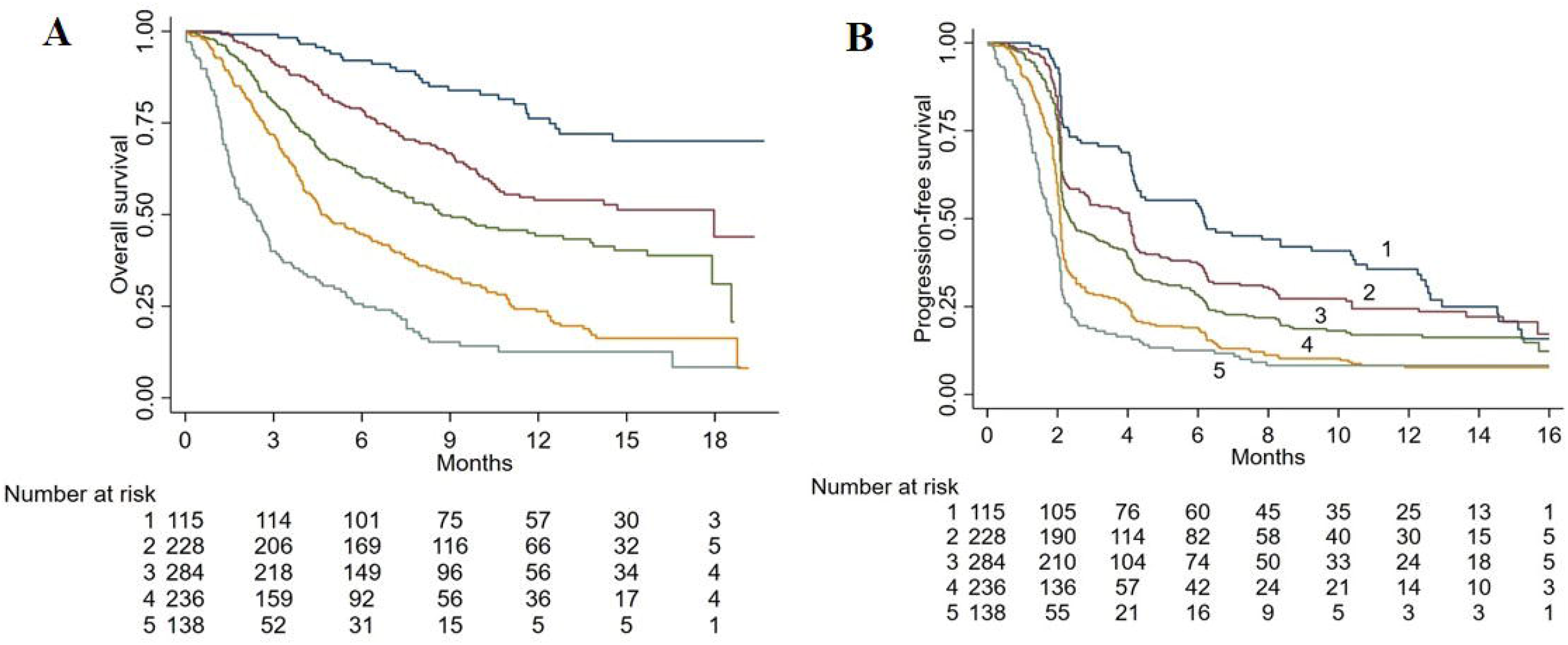
Kaplan Meiers curves for OS (A) and PFS (B) of the prognostic score.

**Figure 2.**
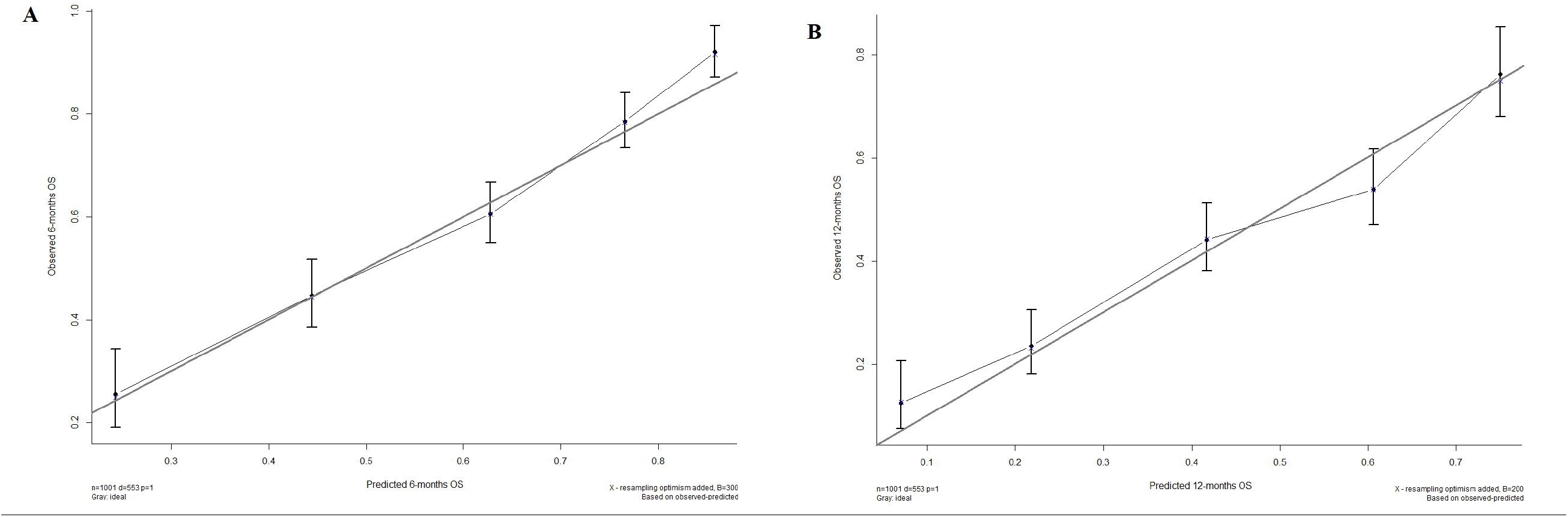
Calibration plot at six months (A) and 12 months (B).

The DCA highlighted a net benefit of the prognostic score when the risk of death at 12 months was higher than 25% (Figure 2S).

## Discussion

Prognostic stratification of patients with aUC might be helpful to clinicians for discussing with patients about their life expectancy and the risk/benefit of available therapeutic options and informing patients’ selection for experimental strategies with novel agents^8^. A few prognostic models based on clinical and laboratory parameters have been tested with immunotherapy^6,8,9,12-14^. By a post-hoc analysis of the Keynote-045 phase III trial^15^, Bellmunt et al. showed that a prognostic model based on either three (ECOG PS > 0, haemoglobin level < 10 g/dL and liver metastases), four (by adding the time from prior chemotherapy <3 months), or five (plus the albumin level lower than 3.5 g/dL) risk factors, was able to accurately stratify the OS of patients treated with pembrolizumab (N = 262) or chemotherapy (N = 267) following first-line platinum-based therapy. Notably, patients treated with pembrolizumab with two, three or four risk factors had similar outcomes, suggesting that novel prognostic models were needed^15^. A prognostic score based on five prognostic factors was then proposed by Sonpavde et al.^6^ through an analysis of 405 aUC post-platinum patients receiving atezolizumab in two phase I/II trials. By including the NLR (with a cut-off of 5), LDH and platelet count, alongside the ECOG PS and liver metastases, the model showed discrimination of survival between low, intermediate and high-risk groups and a slightly superior c-index of 0.692 compared to 0.635 for the 3-factor Bellmunt score^6^. Validation in an independent cohort of patients arising from two phase I/II studies of avelumab (N = 242) and durvalumab (N = 198) was also provided^6^. By a post-hoc analysis of the Italian patient cohort of the SAUL phase IIIB study^7^, including 267 real-world post-platinum aUC patients treated with atezolizumab, we reported that the combination of two laboratory parameters, or the SII (with cut-off >1375) and PD-L1 status, with or without the addition of LDH, discriminated both the OS and PFS of three risk groups (low, intermediate and high risk)^8^.

Regarding the first-line immunotherapy, we initially reported that a combination of NLR (with a cut-off of 5) and LDH predicted both OS and PFS^9^. In this treatment setting, Khaki et al.^16^ developed a prognostic model based on the combination of NLR (with a cut-off of 5) with ECOG PS (≥2), albumin (<3.5 g/dL) and liver metastases. The score was able to stratify the OS of 357 real-world aUC patients receiving first-line ICIs in four risk groups (with 0, 1, 2 and ≥ 3 risk factors). The c-index of 0.68 was slightly superior to the 0.63 of the 2-risk Bajorin’s consisting of the Karnofsky performance status (KPS) (less than 80%) and visceral (lung, liver, or bone) metastases^13^.

With the current analysis of the whole SAUL cohort, we developed a score based on eight independent prognostic factors with different risk weights contributing to the total scoring. This score identified five risk groups with a c-index superior on both OS (0.702 versus 0.672) and PFS (0.694 versus 0.613) compared to Bellmunt’s one. Differently from Bellmunt’s score, the model considered the ECOG PS as ≥ 1 (instead of ≥ 0) and added to the haemoglobin level and liver metastases other two clinical factors, i.e. the bone metastases and concomitant use of steroids, and three laboratory ones, the SII (≥884), LDH (>ULN) and histological subtype (i.e., non-urothelial). Furthermore, each included factor had a different regression coefficient-based weight on the prognostic score. The prognostic models developed with the immunotherapy^6,8,16^ had already added to the Bellmunt score a peripheral blood-derived immune-inflammatory biomarker to the NLR. The negative prognostic value of NLR has been confirmed in several tumour types^17-18^, including the aUC treated with either immunotherapy^6,8,9^ or chemotherapy^19-20^ and other urological cancers treated with immunotherapy^21^. Recently, NLR was indicated as a possible predictive factor for advanced non-small-cell lung cancer treated with single-agent immunotherapy after progression to first-line chemotherapy^22^. In our previous analysis of the Italian SAUL patient cohort^8^, we found that the SII, with the receiving operating curve (ROC)-derived cut-off of 884, performed slightly better than NLR in terms of prognostic accuracy, which was confirmed in the current analysis. One reasonable explanation is that SII incorporates the platelet count, which was included in the Sonpavde’s as a separate factor^6^. Furthermore, the proposed score here encompasses three other baseline factors more likely linked to the effect of the immunotherapy. While bone metastases have already been indicated as an independent adverse prognostic factor in patients with aUC receiving single-agent immunotherapy^23^, concomitant steroids and the non-urothelial histological subtype were not previously studied since these represented exclusion criteria of phase II and III trials with immunotherapy in aUC. Therefore, those could not be identified by Sonpavde’s analysis, which relies on phase I/II studies^6^. Moreover, using concomitant steroids with immunotherapy is a known adverse prognostic factor and has already entered into prognostic models based on real-world patients with other cancer types^24^.

For the above reasons, the current score may be more contemporary and considers parameters that might correlate to the effect of immunotherapy. The large sample size represented by real-world patients, followed up according to the criteria of a phase IIIb trial, is an additional strength of our analysis and mirrors the population of patients clinicians face in their daily clinics. Furthermore, the current score was developed in chemotherapy pretreated patients receiving single-agent ICI and, deliberately, with baseline factors untied from the previous treatment, such as the time elapsed from the last chemotherapy cycle or the best response to the chemotherapy. Thus, the score might remain discriminatory in other treatment settings, like maintenance or novel immunotherapy combinations.

Among the limitations of the current analysis, we acknowledge the lack of external validation, long-term follow-up, and testing of ICIs other than atezolizumab. However, the follow-up was adequate for the second-line setting and did not seem to affect the OS and PFS curves provided by the model. The other two limitations should have been mitigated by the bootstrapping method performed on a large sample size and the similar activity of ICIs shown across clinical trials, respectively. Furthermore, the lack of control treatment does not allow drawing any conclusions regarding the predictive value of the model or its variables.

Nonetheless, the current score provides robust prognostic discrimination of patients homogeneously treated with immunotherapy through baseline affordable and reproducible clinical and laboratory factors, making the current score widely applicable in clinical practice at no additional costs.

It could assist clinicians in discussing the prognosis with patients who are offered immunotherapy in clinical practice and used to stratify patients in clinical trials with novel immunotherapy combinations. For instance, a hypothesis generated by the model to be tested in clinical trials is that patients belonging to group 1 might benefit from immunotherapy only, those in group 2,3,4 of immunotherapy combination, whilst it is pretty unlikely that patients in group 5 will take advantage of immunotherapy even if combined with novel agents. Moreover, a better prognostic tool could direct clinicians to different treatment strategies according to the different prognostic stratification (e.g. treatment beyond progression or change of treatment).

More sophisticated prognostic models could provide better prognostic discrimination, like that developed by Nasser et al. on 62 aUC patients treated with ICIs who had targeted tumour sequencing. It was based on NLR, visceral metastases and single-nucleotide variants, and the c-index was 0.90^12^. However, the affordability and reproducibility of prognostic models embodying genomic testing must always be pondered.

In conclusion, we proposed a 5-classes prognostic model contemporary to immunotherapy based on eight affordable clinical or laboratory factors. This prognostic score could be adopted in clinical practice to inform patients about prognosis with immunotherapy and assess the benefit of novel immunotherapy combinations in clinical trials.

## Data Availability

Data availability is subject to a written request to Roche.

## Figure legend

**Figure 1S.**Kaplan Meiers curves for OS (A) and PFS (B) of the Bellmunt score.

**Figure 2S**.Decision curve analysis (DCA) for the net benefit of the prognostic score based on 12-month overall survival.

## Acknowledgements

All authors would like to thank F. Hoffmann-La Roche for the approval of this subanalysis project and sharing of the dataset of the original study, in particular Cinzia Astolfi for her technical support.

We thank the Meet-URO Italian Network For Research In Urologic-Oncology for supporting the analysis’s planning and results’ interpretation. Dr Rebuzzi and Dr Fornarini would like to thank the Italian Ministry of Health (Ricerca Corrente 2018-2021 grants) for financially supporting their research on identifying prognostic and predictive markers for patients with genitourinary tumours. Dr Rescigno’s work is funded by Prostate Cancer Foundation through a PCF YI award and by the FPRC 5 PER MILLE - Ministero della Salute 2017 - PTCRC SEE PROS ONCOLOGIA and “Italian Ministry of Health, Ricerca Corrente 2022”.

## Author contributions

Study concept and design, G.F., S.E.R., A.S. and G.L.B.; G.F. and S.E.R. contributed equally as first authors; G.L.B. and A.S. contributed equally as senior authors;. Acquisition and curation of data, all authors; statistical analysis, A.S.; interpretation of data, G.F., S.E.R., A.S., G.L.B., S.B. and P.R.; drafting of the manuscript, S.E.R., A.S., G.L.B. and P.R.; critical revision of the manuscript for important intellectual content: G.F., S.E.R., A.S., G.L.B., S.B. and P.R.; supervision, S.E.R., G.L.B. and G.F. All authors have read and agree to the published version of the manuscript.

