## Supplemental Table 1 for "A novel immunotherapy prognostic score for patients with pretreated advanced urinary tract carcinoma from the subgroup analysis of the SAUL study. The urInary TrAct CArcinoma score (ITACA)"

**Table 1S. Correlation of the prognostic score with PFS.**

| **Risk score groups (original score)** | **N (total subgroups)** | **mPFS (95% CI)** | **HR (95% CI)** | **p value** | **c-index** |
| --- | --- | --- | --- | --- | --- |
| 1 (0-1) | 115 (4+111) | 6.18 (4.23-9.23) | 1.00 (ref) |  | 0.694  (95% CI: 0.673-0.715 |
| 2 (2-3) | 228 (53+175) | 4.04 (2.79-4.17) | 1.33 (1.02-1.73) | 0.038 |  |
| 3 (4-5) | 284 (111+173) | 2.33 (2.14-3.12) | 1.66 (1.28-2.15) | <0.001 |  |
| 4 (6-7) | 236 (118+118) | 2.07 (2.00-2.14) | 2.45 (1.89-3.17) | <0.001 |  |
| 5 (8+) | 138 (59+51+19+9) | 1.81 (1.51-2.00) | 3.41 (2.56-4.54) | <0.001 |  |

Abbreviations: CI, confidence interval; HR, hazard ratio; mPFS, median progression-free survival; N, number of patients; PFS, progression-free survival; ref, reference.
