## Supplemental Table 2 for "A novel immunotherapy prognostic score for patients with pretreated advanced urinary tract carcinoma from the subgroup analysis of the SAUL study. The urInary TrAct CArcinoma score (ITACA)"

**Table 2S. Correlation between the Bellmunt score and OS**

| **Bellmunt score** | **N (total subgroups)** | **mOS (95% CI)** | **HR (95% CI)** | **p value** | **c-index** |
| --- | --- | --- | --- | --- | --- |
| 0 | 314 (31.4) | 17.9 (13.8-21.0) | 1.00 (ref) |  | 0.672  (95% CI: 0.643-0.701) |
| 1 | 404 (40.4) | 8.48 (7.46-10.58) | 1.85 (1.48-2.31) | 0.001 |  |
| 2 | 244 (24.4) | 3.88 (3.12-4.53) | 3.95 (3.13-4.99) | <0.001 |  |
| 3 | 39 (3.9) | 1.97 (1.18-2.79) | 6.56 (4.40-9.77) | <0.001 |  |

Abbreviations: CI, confidence interval; HR, hazard ratio; mOS, median overall survival; N, number of patients; NR, not reached; OS, overall survival; ref, reference.
