## Supplemental Table 3 for "A novel immunotherapy prognostic score for patients with pretreated advanced urinary tract carcinoma from the subgroup analysis of the SAUL study. The urInary TrAct CArcinoma score (ITACA)"

**Table 3S. Correlation between the Bellmunt score and PFS.**

| **Bellmunt score** | **N (total subgroups)** | **mPFS (95% CI)** | **HR (95% CI)** | **p value** | **c-index** |
| --- | --- | --- | --- | --- | --- |
| 0 | 314 (31.4) | 4.11 (3.45-4.34) | 1.00 (ref) |  | 0.613  (95% CI:  0.589-0.637) |
| 1 | 404 (40.4) | 2.30 (2.14-2.79) | 1.33 (1.12-1.58) | 0.001 |  |
| 2 | 244 (24.4) | 2.07 (1.94-2.10) | 2.07 (1.72-2.51) | <0.001 |  |
| 3 | 39 (3.9) | 1.31 (1.15-1.84) | 4.11 (2.88-5.85) | <0.001 |  |

Abbreviations: CI, confidence interval; HR, hazard ratio; mPFS, median progression-free survival; N, number of patients; PFS, progression-free survival; ref, reference.
