## Supplementary figures and images for "A novel immunotherapy prognostic score for patients with pretreated advanced urinary tract carcinoma from the subgroup analysis of the SAUL study. The urInary TrAct CArcinoma score (ITACA)"

### Supplemental Figure 1

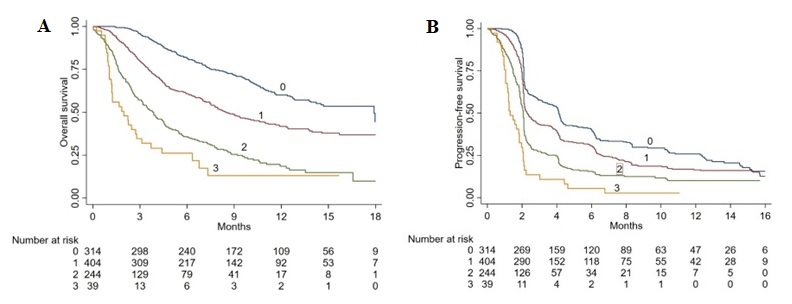

### Supplemental Figure 2

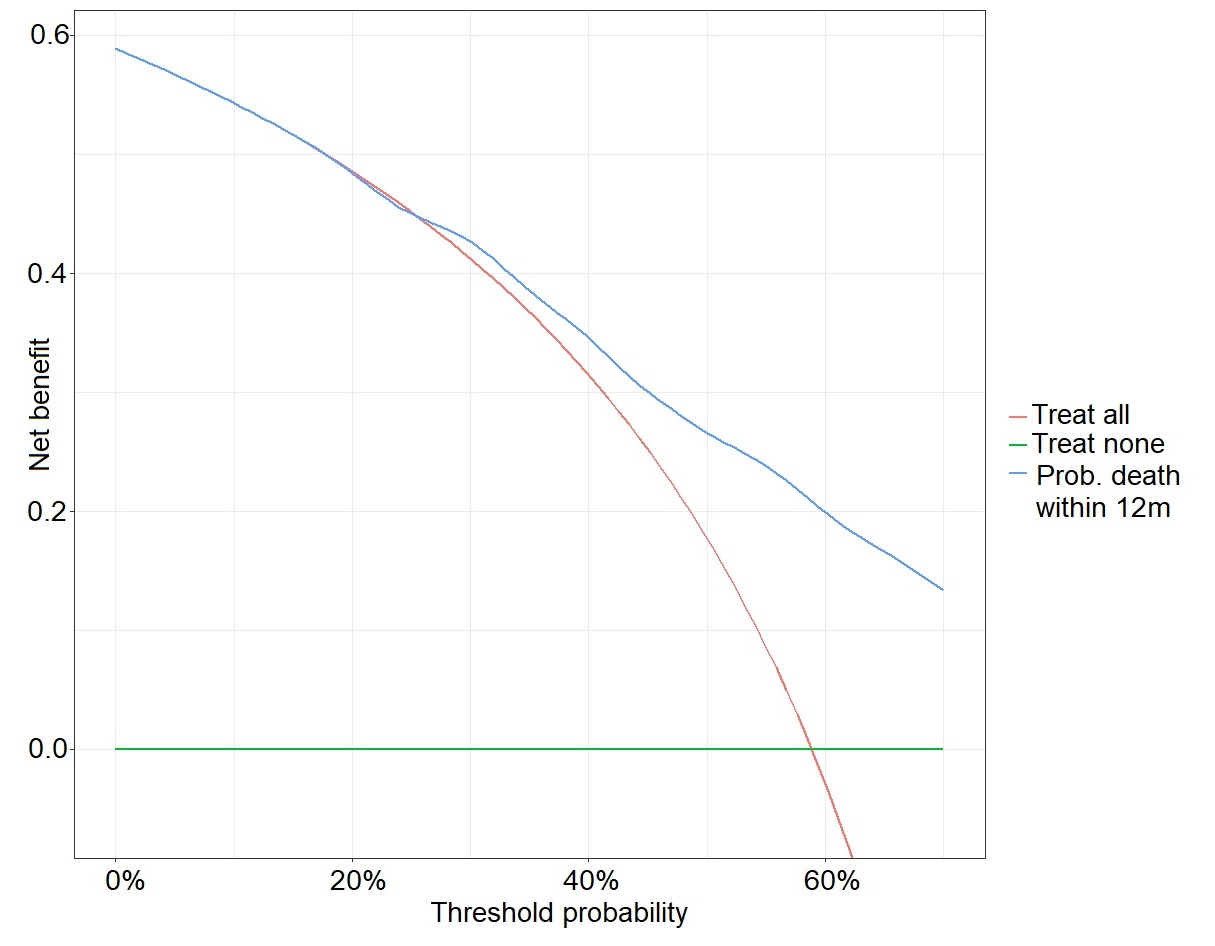
